# STRESS, ANXIETY AND PTSD PREVALENCE AMONG UKRAINIANS GREW DRAMATICALLY DURING THE FIRST YEAR OF RUSSIAN INVASION: RESULTS OF NATIONWIDE SURVEY

**DOI:** 10.1101/2023.06.24.23291803

**Authors:** Oleh Lushchak, Mariana Velykodna, Svitlana Bolman, Olha Strilbytska, Vladyslav Berezovskyi, Kenneth B. Storey

**Affiliations:** Department of Biochemistry and Biotechnology, Precarpathian National University, Ivano-Frankivsk, Ukraine (Prof O Lushchak PhD, O Strilbytska PhD, V Berezovskyi); Research and Development University, Ivano-Frankivsk, Ukraine (O Lushchak, S Bolman); Kryvyi Rih State Pedagogical University, Kryvyi Rih, Ukraine (Prof M Velykodna MD, PhD); National Psychological Association, Kyiv, Ukraine (M Velykodna, S Bolman); Carleton University, Ottawa, Canada (Prof K Storey PhD)

## Abstract

**Background:** In February 2022 the Russian federation started a new invasion of Ukraine as an escalation of the ongoing war since 2014. After nine years of war and the COVID-19 pandemic, the mental health state of Ukrainians requires systematic monitoring and relevant action.

**Methods:** This study was designed as an online survey arranged in the 9-12 months after the start of the new invasion of Ukraine and includes sociodemographic data collection, evaluation of stress intensity by PSS-10, anxiety with GAD-7, and symptoms of posttraumatic stress disorder with PCL-5. The sample size of 3173 Ukrainians consisted of 1954 (61.6%) respondents that were not displaced persons (NDPs), 505 (15.9%) internally displaced persons within Ukraine (IDPs), and 714 (22.5%) refugees that left Ukraine.

**Findings:** Moderate and high stress was prevalent among 64.7% and 15.5% of NDPs, 64.4% and 21.6% of IDPs, and 68.2% and 25.2% of refugees, respectively. Moderate and severe anxiety was prevalent among 25.6% and 19.0% of NDPs, 25.7% and 23.4% of IDPs, and 26.2% and 25.8% of refugees. High levels of PTSD (33 and higher) were prevalent among 32.8% of NDPs, 39.4% of IDPs, and 47.2% of refugees. DSM-V criteria for PTSD diagnosis was met by 50.8% of NDPs, 55.4% of IDPs, and 62.2% of refugees.

**Interpretations:** The lowest stress, anxiety, and PTSD severity was observed among NDPs, with significantly higher levels among IDPs and the highest among refugees. Being forcibly displaced from the previous living area and, especially, entering a new cultural environment significantly contributes to the mental health issues caused by war exposure, particularly for people who have directly witnessed the results of war.

**Fundings:** Ministry of Education and Science of Ukraine

**Research in context:** *Evidence before this study:* Previously published evidence suggested an increase and specificity of stress, anxiety, and PTSD prevalence among Ukrainians by August 2022. Some research provided a data comparison between IDPs and refugees. However, to date, no study has reported a comparison of three groups of Ukrainians: NDPs, IDPs, and refugees abroad.

*Added value of this study:* This is the first study that provides evidence of the mental health state of Ukrainians after 9-12 months of the Russian invasion in 2022. Furthermore, the research presented was designed as a nationwide survey involving three groups for comparison: NDPs and IDPs within Ukraine, and refugees in other countries. The results of the study show a significant difference between these groups in stress, anxiety, and prevalence of PTSD symptoms.

*Implications of all available evidence:* The present study contributes to the comprehension of the dynamics in mental health of Ukrainians in response to the war. This data will be helpful both in mental health strategy development by governmental and international policy and in the local clinical work of mental health professionals working with Ukrainians.

## Introduction

In February 2022 the Russian federation started a new invasion of Ukraine as an escalation of the war started in 2014.^1^ Ukrainian people faced numerous issues caused by war exposure and witnessing, from the danger of death and physical injury to various symptoms of mental trauma.^2–4^ By November 2022, when our research started, the Office of the UN High Commissioner for Human Rights (OHCHR) had reported 6,655 killed and 10,368 injured people, including 419 and 769 children, respectively.^5^ The scale of this war and the Russian narrative support for invasion allowed researchers and politicians to categorize this as genocide and to predict its local and global consequences.^6–9^

The first survey report provided evidence of war effects on the mental health of the Ukrainian population as of March 2022, when researchers reported that 53% of Ukrainian adults experienced severe mental distress, 54% with anxiety, and 47% with depression.^4^ In April 2022, 30.8% of surveyed adults displaced within Ukraine and abroad met the criteria for elevated risk for PTSD without a significant difference between groups.^2^ Six months after the invasion, 26% of parents located in Ukraine had PTSD, and 15% had developed complex posttraumatic stress disorder (CPTSD).^3^ By May-August 2022, researchers reported anxiety symptoms among 51% of adult Ukrainian refugees, distress among 23%, and severe distress among 41%.^10^ Further war-related psychological symptoms, including CPTSD and prolonged grief disorder (PGD), are expected in the coming months, years, decades, and future generations.^11^

Researchers suggested that the previous invasion in 2014 along with the COVID-19 pandemic have significantly contributed to the currently observed mental health issues.^12–14^ The prevalence of stress symptoms in the general population of Ukrainian adults increased from 45% in 2012 to 50% in 2016.^15^ Adolescents living in a war-torn region of Eastern Ukraine in 2016-2017 had significantly increased risks for PTSD, severe anxiety, and depression than their peers from other regions.^16^ In 2017, Global Burden of Disease reported 14% of mental disorders prevalent in the general Ukrainian population, including 3% of anxiety and 5% of depression.^17^ Therefore, ongoing monitoring of current and further dynamics in the mental health of Ukrainians is needed.

The present study aimed to investigate the state of mental health among Ukrainians after 9-12 months of invasion to assess the levels and changes in their stress, anxiety, and PTSD prevalence in three groups: not displaced Ukrainians (NDPs), IDPs, and refugees abroad. For this purpose, we elaborated and conducted a complex nationwide survey among people within Ukraine and abroad. We compare and discuss our results with those from previous studies.

## METHODS

### Survey elaboration

An online survey was considered as the most appropriate tool for this study to reach people across the country and abroad. The survey included gathering sociodemographic data (age, sex, education, current and prior location, social and mental health status) and three validated self-report questionnaires to measure stress, anxiety, and PTSD levels. All questionnaires had Ukrainian versions adapted and validated by earlier studies.^18–20^

### Stress

Stress was assessed using the Perceived Stress Scale (PSS-10), a ten-item questionnaire with a total score of 0 to 40.^21^ The respondent was asked to assess the frequency of the listed feelings and thoughts observed during the past month using the standard Likert scale from ‘never’ (0) to ‘very often’ (4). Summed scores from 0-13 were considered low stress, 14-26 as moderate, and 27–40 as high perceived stress.^20^ Cronbach’s alpha of PSS-10 was estimated at 0·881.

### Anxiety

Anxiety was assessed using the GAD-7 questionnaire that estimates generalized anxiety disorder.^22^ This instrument was validated for use in primary care and general populations.^23^ Besides, the GAD-7 was extensively used among Ukrainians as a reliable instrument, which allows data comparison with previous studies.^16^ This self-report seven-item anxiety questionnaire determines the mental health state of an individual during the previous two weeks. The respondent had to assess the degree to which he or she felt nervous, anxious, unable to control worrying, easily irritable, and afraid that something might happen as ‘not at all’, for ‘several days’, for ‘more than half the days’ and for ‘nearly every day’. Total scores from 0 to 21 were categorized as follows: up to 5 no or minimal anxiety, up to 10 mild anxiety, up to 15 as moderate anxiety, and 16 and above as severe anxiety. A cutoff score of 10 was recommended to identify people who might have an anxiety disorder that requires further diagnostics.^23^ Cronbach’s alpha of GAD-7 for this study was calculated as 0·889.

### Posttraumatic stress disorder

Posttraumatic stress disorder was assessed using the PTSD Checklist for DSM-5 (PCL-5) developed for screening PTSD and monitoring its symptom changes.^24^ PCL-5 is a self-assessment questionnaire with 20 items scored on a Likert scale from 0 (‘Not at all’) to 4 (‘Extremely’). The total PTSD symptom severity score (from 0 to 80) can be obtained by summing the scores of all items. A cutoff of 33 is considered indicative of PTSD among samples, including Ukrainian adults.^18^ Another way to interpret PCL-5 results is to calculate its separate Criteria: A - trauma exposure or witnessing; B - intrusion symptoms; C - avoidance; D - negative alterations in cognition and mood; E - alterations in arousal and reactivity. The DSM-5 diagnostic rule for PTSD diagnosis requires that at least 1 B item, 1 C item, 2 D items, and 2 E items should be rated as 2 (‘Moderately’) or higher. Cronbach’s alpha of the total score of PCL-5 in this study was 0·944. Reliability of the other PCL-5 scales was also satisfactory (Criterion B, 0·873; Criterion C, 0·784; Criterion D, 0·885; Criterion E, 0·816).

### Procedure

The survey responses were gathered via filling out of Google Forms along with written consent to participate in the study. Participation was voluntary and confidential. The survey was conducted among adults within Ukraine, internally displaced people, and refugees from November 2022 to January 2023, i.e. a time of 9 to 12 months after the invasion. During this period, there were constant missile attacks throughout Ukraine, which caused long blackouts in almost all regions of the country. It was a period when many people had no electricity and heating for days or even weeks.

### Data analysis

Data analysis was performed in SPSS Statistics 22 software (IBM, Armonk, NY, USA). The results presented were extracted with descriptive statistics for data presentation, Cronbach’s alpha coefficients for reliability calculation, Shapiro-Wilk to test the normality of the distribution, Kruskal-Wallis test for ordinary data with further pairwise comparison using the Mann-Whitney U-test, and Pearson Chi-square test and Cramer’s V test for frequencies. Figures were generated in Prism GraphPad (GraphPad Software, MA, USA)

## RESULTS

### Survey participants

We gathered 3173 complete responses from Ukrainian adults 18-92 years old, with 1954 (61.6%) participants that stayed within their permanent location (i.e., were NDPs: not displaced persons), 505 (15.9%) were internally displaced within Ukraine (IDPs: internally displaced persons), and 714 (22.5%) had fled abroad (as refugees). Since the general population of Ukraine was about 43.8 million in 2021, according to information from The World Bank, the sample size corresponds to a confidence level of 99% with a 2.3% margin of error. The geographical distribution of respondents before the invasion is provided in figure 1 and shows that we succeeded in covering all regions of the country. This dataset and number of respondents allowed us to define mental health not only in NDPs but also evaluate the effects on INPs or those that moved abroad as refugees (table 1).

**Figure 1.**
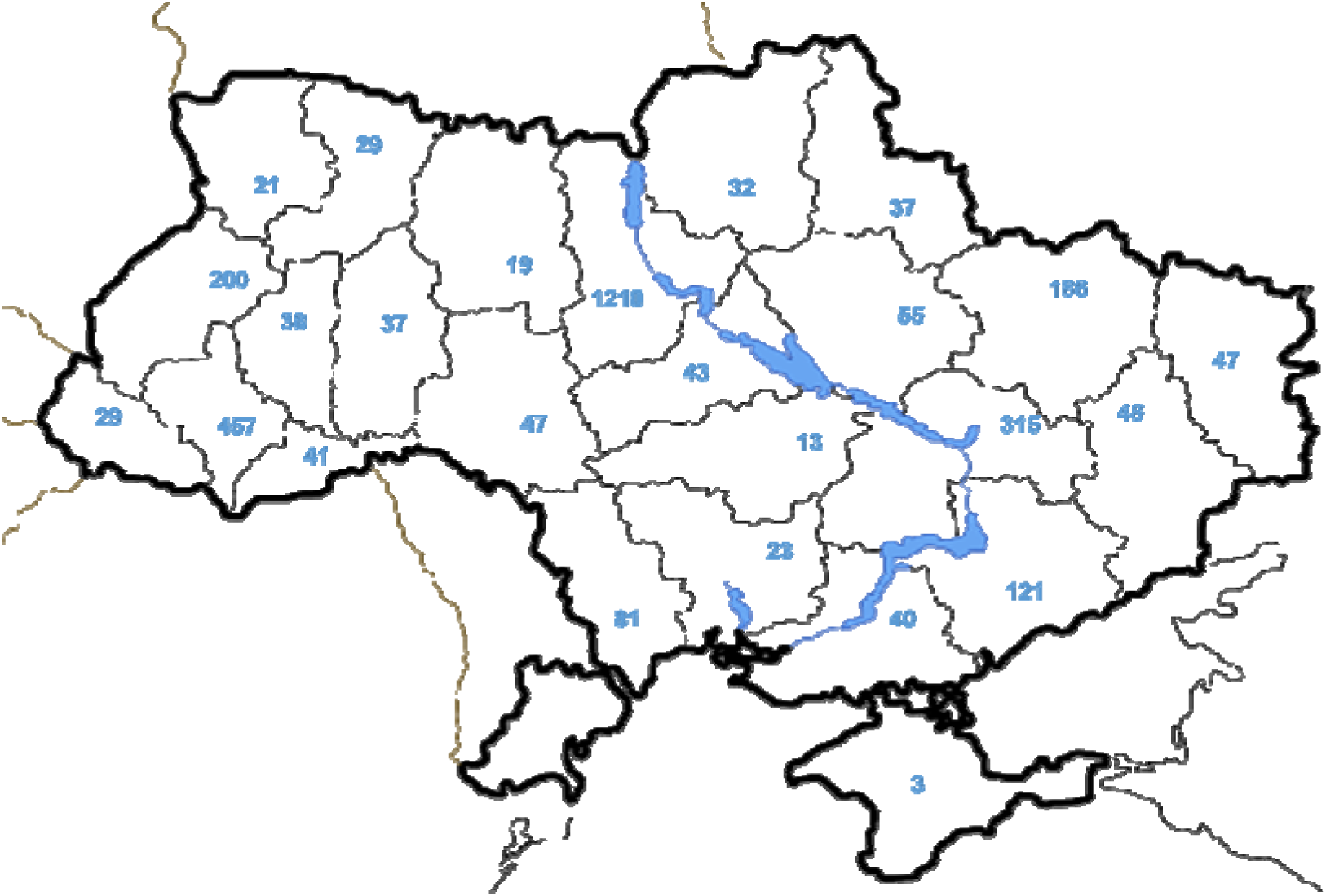
Geographical distribution of respondents before 24 February 2022.

**Table 1.** Sociodemographic group description and comparison

|  | NDUs | IDPs | Refugees | Cramer's V | P value |
| --- | --- | --- | --- | --- | --- |
| Sex |  |  |  |  |  |
| Female | 1625 (83.2%) | 437(86.5%) | 678(95.0%) | 0.1395 | <0.0001 |
| Male | 329(16.8%) | 68(13.5%) | 36(5.0%) |  |  |
| Social status |  |  |  |  |  |
| Not married | 607(31.1%) | 127(25.1%) | 189 (26.5%) | 0.0871 | <0.0001 |
| Married | 1153(58.9%) | 323(64.0%) | 393 (55.0%) |  |  |
| Divorced | 187(9.6%) | 55(10.9) | 131 (18.4%) |  |  |
| Other | 7(0.4%) | 0 (0.0%) | 1 (0.1 %) |  |  |
| Kids |  |  |  |  |  |
| No children | 824 (42.2%) | 165 (32.7%) | 216 (30.3%) | 0.1004 | <0.0001 |
| 1 child | 545 (27.9%) | 195 (38.6%) | 269 (37.7%) |  |  |
| 2 children | 479 (24.5%) | 130 (25.7%) | 172 (24.1%) |  |  |
| 3 children | 88 (4.5%) | 11 (2.2%) | 50 (7.0%) |  |  |
| 4+ children | 17 (0.9%) | 4 (0.8%) | 7 (1.0%) |  |  |
| Highest education level |  |  |  |  |  |
| Secondary school | 60 (3.1%) | 9 (1.8%) | 13 (1.8%) | 0.0361 | 0.0817 |
| High school | 118 (5.8%) | 27 (5.3%) | 29 (4.1%) |  |  |
| Higher institution | 1780 (91.1%) | 469 (92.9%) | 672 (94.1%) |  |  |
| Have been in zones of active hostilities |  |  |  |  |  |
| As a civilian | 218 (11.2%) | 203 (39.9%) | 164 (23%) | 0.2736 | <0.0001 |
| As a military | 67 (3.4%) | 11 (2.2%) | 3 (0.4%) | 0.0781 | <0.0001 |
| None | 1680 (85.4%) | 294 (57.9%) | 547 (76.6%) | 0.2472 | <0.0001 |
| Age |  |  |  |  |  |
| Minimum >> Maximum | 18 >> 80 | 18 >> 72 | 18 >> 92 |  |  |
| Average \| St- deviation | 37.13 \| 11.19 | 38.53 \| 9.51 | 38.44 \| 9.35 |  |  |
| Age distribution |  |  |  |  |  |
| 18-25 years | 301 (15.4%) | 35 (6.9%) | 4 (6.3%) | 0.1089 | <0.0001 |
| 26-35 years | 578 (29.6%) | 157 (31%) | 214 (30%) |  |  |
| 36-45 years | 640 (32.8%) | 208 (41.2%) | 319 (44.7%) |  |  |
| 46-59 years | 372 (19%) | 90 (17.8%) | 116 (16.2%) |  |  |
| 60+ years | 63 (3.2%) | 15 (3%) | 20 (2.8%) |  |  |
| Total | 1954 (61.6%) | 505 (15.9%) | 714 (22.5%) |  |  |

Females were the largest numbers in all three groups, representing 95% among refugees. Those aged 26 years and older or who had children were more often displaced within Ukraine or abroad. Married people were more likely to flee within Ukraine, whereas divorced people generally moved abroad and single persons most often stayed in their permanent residence. Direct exposure or witnessing the effects of war forced civilians to flee from their areas of residence, whereas war-exposed persons with current or past military experience tended to stay in their permanent locations.

### Mental health in general

In Table 2, we present the self-reported mental health issues that the respondents had diagnosed in the previous periods. In sum, 20.8% of NDPs, 21% of IDPs, and 27.7% of refugees confirmed diagnosed mental health issues, including specific disorders. Refugees reported previously diagnosed mental health issues most often (in 27.7% of cases, p=0·0006), especially anxiety disorders (p=0·0027) and depression (p=0·0066). In addition, refugees were more likely to receive mental health care regularly (p<0·001).

**Table 2.** Mental health issues within groups before and after the 2022 invasion

| Group | NDPs | IDPs | Refugees | Cramer's V | P value |
| --- | --- | --- | --- | --- | --- |
| **Previously diagnosed mental health issues** |  |  |  |  |  |
| Anxiety disorders | 274 (14%) | 69 (13.7%) | 137 (19.2%) | 0.0612 | 0.0027 |
| Depression | 243 (12.4%) | 64 (12.7%) | 122 (17.1%) | 0.0563 | 0.0066 |
| PTSD | 48 (2.5%) | 14 (12.8%) | 19 (2.7%) | 0.008 | 0.9029 |
| Other | 109 (5.6%) | 25 (5%) | 40 (5.6%) | 0.0102 | 0.8479 |
| None | 1547 (79.2%) | 399 (79%) | 516 (72.3%) | 0.0688 | 0.0006 |
| **Received mental health care** |  |  |  |  |  |
| Regularly | 143 (7.3%) | 39 (7.7%) | 94 (13.2%) | 0.1029 | <0.0001 |
| Sometimes | 335 (17.1%) | 93 (18.4%) | 194 (27.2%) |  |  |
| No | 1476 (75.5%) | 373 (73.9) | 426 (59.7%) |  |  |
| **Prevalence of stress, anxiety or PTSD** |  |  |  |  |  |
| No | 164 (8.4%) | 31 (6.2%) | 34 (4.8%) | 0.06 | 0.0033 |
| Yes | 1785 (91.6%) | 472 (93.8%) | 680 (95.2%) |  |  |

To evaluate the current mental health of the studied groups in general, we calculated the percentage of the respondents who reached a moderate or severe level of at least one of the measured variables (table 2). Following table 2, only 7.2% of the respondents reported no or mild stress, anxiety, and PTSD levels within the sample. Others had mental health issues as shown by one or more variable. Refugees reported mental health issues in wartime more often (p=0·0033).

### Stress

Representatives of all groups showed high levels of stress (figure 2). Only 16.3% of NDPs had low levels of stress, whereas 68.2% had moderate stress and 15.5% perceived high stress, respectively (figure 2A). Leaving homes significantly increased the percentage of individuals with high perceived stress to 21.3% for IDPs and 25.5% for refugees. Evaluation of the total score obtained by summing points for every question showed the difference between groups (figure 2A). The perceived stress prevalence differed significantly among representatives of the three groups (figure 2B). People who had to flee from their prior residence within the country, i.e., IDPs (M=20·97, SD=6·86), had higher stress levels than NDPs (M=19·88, SD=6·49), whereas the refugees had the highest stress levels (M=22·1, SD=6·37).

**Figure 2.**
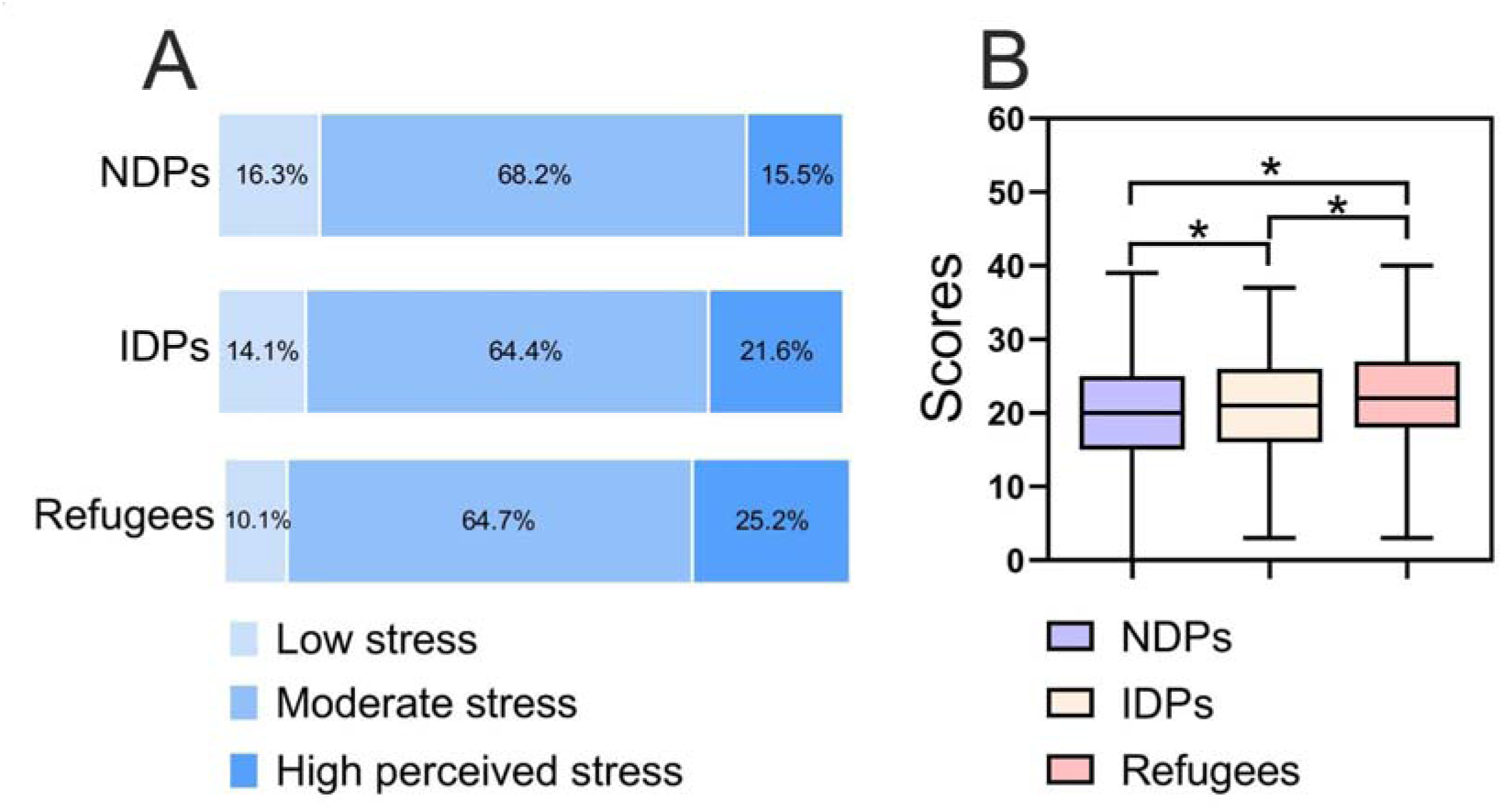
The prevalence of perceived stress among the respondents of NDPs, IDPs, and refugee groups. A - percentage of respondents with low (0-13), moderate (14-26), and highly perceived stress (26-40) according to the PSS-10 questionnaire; B - total scores from the questionnaire represented as median, interquartile range from 25 to 75%, and maximum and minimum values shown by whiskers. *Significant difference between groups with p<0·05 compared by Kruskal-Wallis test.

### Anxiety

Within the sample, the prevalence of moderate anxiety was 25.6% to 26.2%, and severe anxiety was inherent to 19-25.6% of the respondents (figure 3A). Anxiety levels also differed significantly between the three studied groups (figure 3B), reflecting the highest levels within the group of refugees (M=10·97, SD=5·67) and the lowest among NDPs (M=9·85, SD=5·41). However, there was no significant difference between the refugees and IDPs (M=10·57, SD=5·7). According to GAD-7 criteria, people with moderate and severe anxiety levels (between 44.6% and 62% in different groups) require further screening.

**Figure 3.**
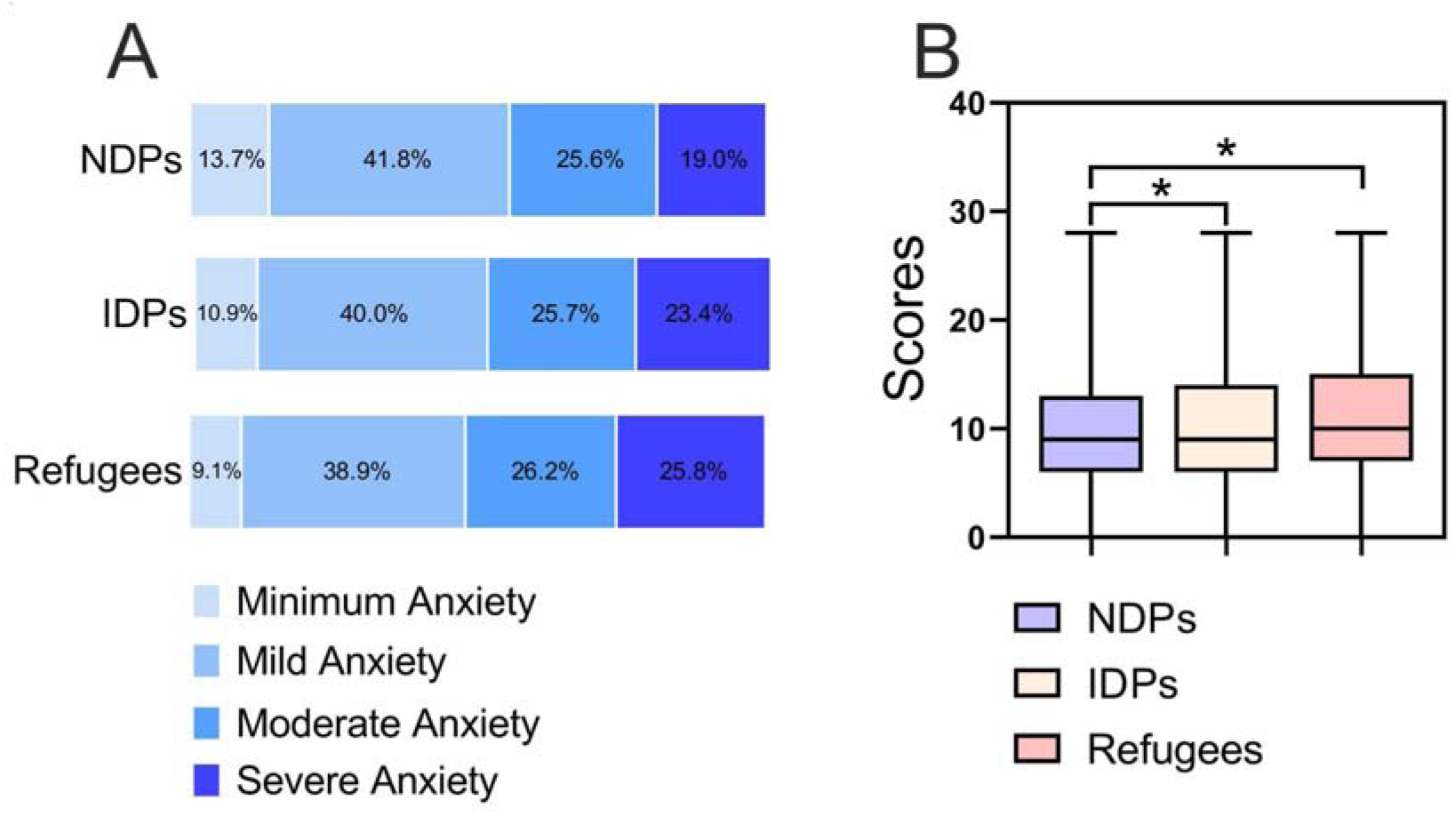
The prevalence of anxiety among respondents of the three groups. A - percentage of respondents with minimum (0-4), mild (5-9), moderate (10-14), or severe anxiety (15-21) according to the GAD-7 questionary. B - total scores from the questionnaire represented by the median, interquartile range, and maximum and minimum of boxplots. *Significant difference between groups with p<0·05 compared by a Kruskal-Wallis test.

### Posttraumatic stress disorder

As PCL-5 has two methods of interpretation, we used both to evaluate PTSD levels within the sample and separate groups. First, we evaluated PTSD levels by identifying people with a 33 score or higher (figure 4A) and found that 32.8% to 47.2% of group respondents had PTSD by this criterion. Group comparison showed that the refugees (M = 31.97, SD = 16.25) had significantly higher and the NPDs (M=26·87, SD=15·98) significantly lower anxiety levels than the IDPs (M=29·52, SD=16·51; figure 4C). Second, we used the formula of PTSD symptoms manifestation sufficient for PTSD diagnosis relevant to DSM-V (figure 4B) and found that 30.8% to 62.2% of surveyed people reached the criteria for PTSD.

**Figure 4.**
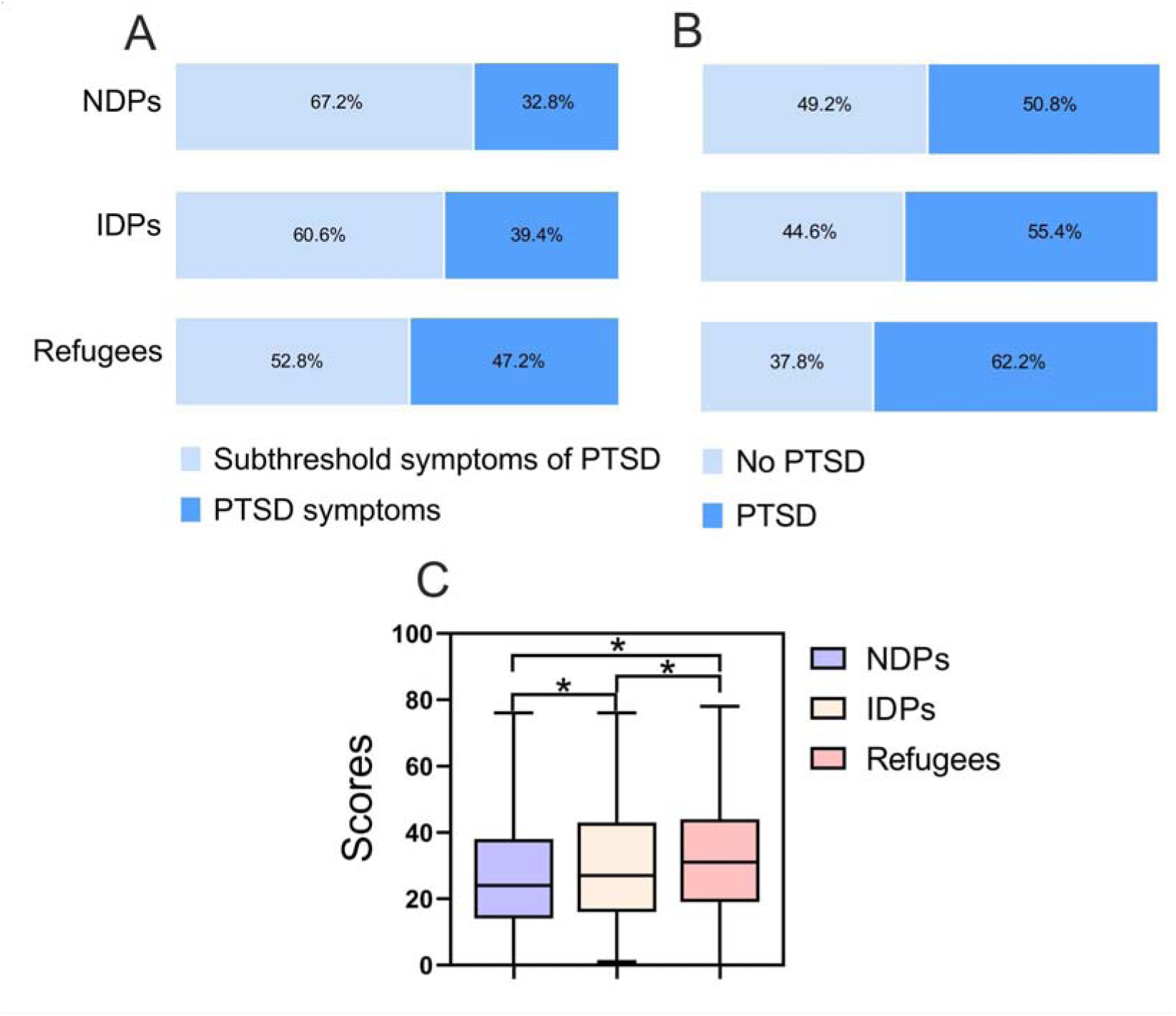
The prevalence of posttraumatic stress disorder among the respondents of three groups. A - percentage of respondents without (0-33) and severe PTSD (33-80) according to the PCL-5 questionary; B - percentage of respondents without and severe PTSD distinguished by criteria; C - total scores obtained within questionary shown by boxplots with representation of median, interquartile range, and maximum and minimum. *Significant difference between groups with p<0·05 as determined by a Kruskal-Wallis test.

Group comparison by the scores of the separate criteria of PCL-5 showed that NDPs had the lowest levels by all variables, whereas IDPs and refugees ranged from no difference to significant difference between groups (figure 5). Intrusion symptoms are significantly higher and equally prevalent among IDPs, and refugees compared to NDPs (figure 5A).

**Figure 5.**
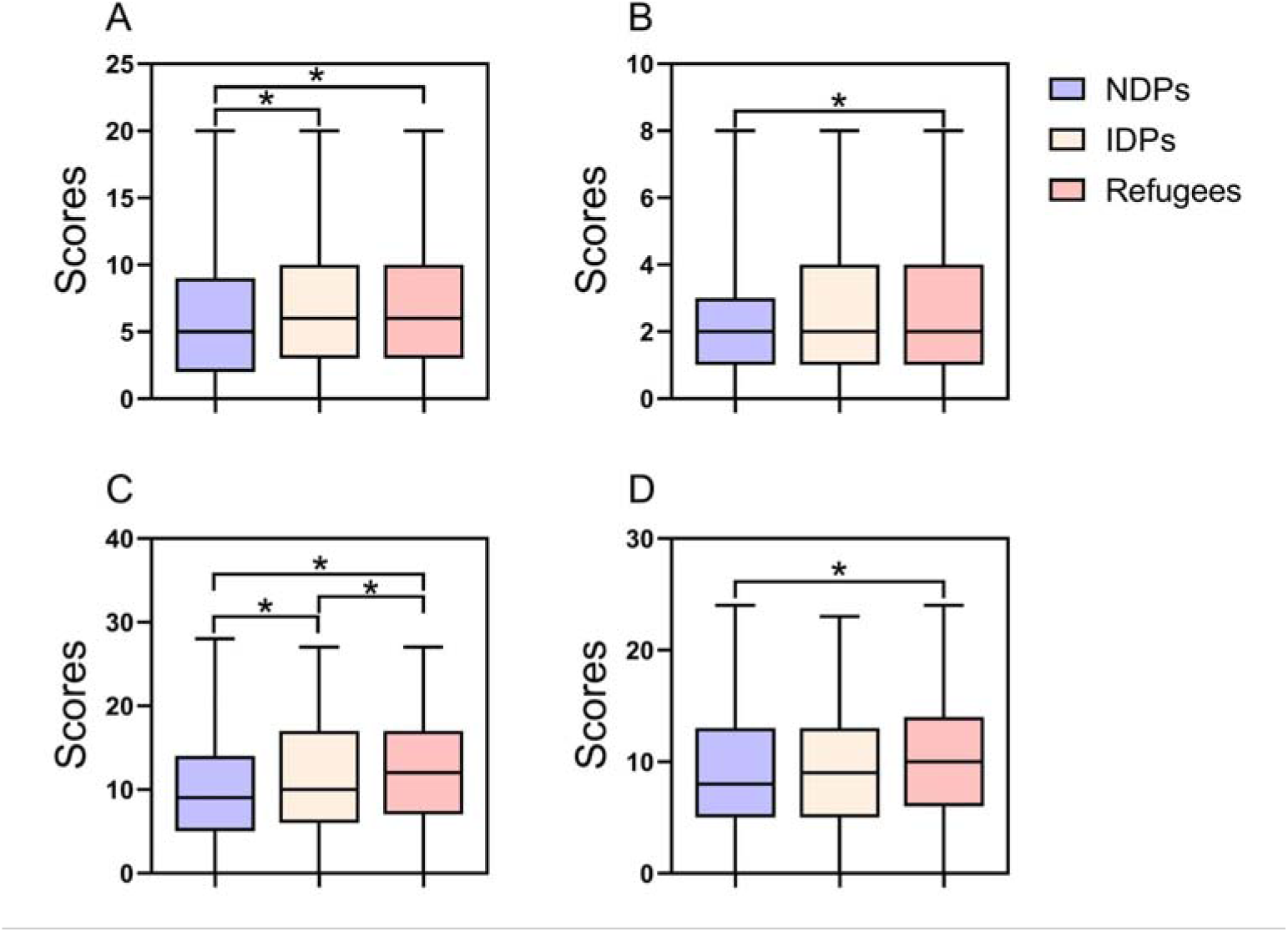
Group comparison by scorers of the PCL-5 criteria. A - Criterion B, B - Criterion C, C - Criterion D, and D - Criterion E. Boxplots represent total scores by each criterion as median, interquartile range from 25 to 75%, and maximum and minimum values shown by whiskers. *Significant difference between groups with p<0·05 compared by a Kruskal-Wallis test.

Avoidance behavior appeared more inherent to refugees than to NDPs whereas IDPs showed no significant difference from the two other groups (figure 5B). As for negative alterations in cognition and mood, there was a significant difference between all three groups, with an increase in this variable from NDPs to IDPs and refugees (figure 5C). Finally, alterations in arousal and reactivity had the highest levels among the refugees compared to NDPs, whereas the IDPs showed no significant differences with other groups (figure 5D).

## Discussion

Foremost, the present data suggest that after 9-12 months of the 2022 Russian invasion of Ukraine, the prevalence of stress, anxiety, and PTSD symptoms among Ukrainian adults reached dramatically high levels. We found that 93% of surveyed people had faced at least one of the measured mental health issues at moderate or severe levels. In comparison, only 20.8-27.7% of respondents reported diagnosed mental health issues before the invasion. This finding might reveal both the severity of ongoing life in wartime and the lack of adequate mental health care and psychological assistance for Ukrainian people suffering from the war.^25^

The current results also show that Ukrainian people who could stay at home (i.e., in their prior place of living) during wartime had significantly lower levels of stress, anxiety, and PTSD compared to those who were forced to flee to other areas. Moreover, the respondents who moved abroad felt even worse than the IDPs by all measured variables except three of the PTSD scales. This allows us to suggest that being forcibly displaced from their previous living area and, especially to a new cultural and social environment, places additional stress and challenges that people must adjust to,^10^ thereby contributing to the mental health issues caused by war exposure or witnessing, compounding previously developed issues.^12^

The current results also contribute to the long-term mental health assessment of Ukrainians, presenting the dynamics of stress, anxiety, and PTSD symptoms prevalence before and after the 2022 Russian invasion of Ukraine. For instance, the prevalence of stress among Ukrainians was 45% in 2012 and 50% in 2016 whereas by March 2022,^15^ stress prevalence increased to 53%.^4^ The prevalence of anxiety in the general population of Ukrainian adults was 54% in March 2022 and 46% in April 2022.^4, 9^ In our study, conducted during constant missile attacks and long blackouts, we obtained 80% moderate and severe stress prevalence and 45% moderate and severe anxiety prevalence among NDPs.

Regarding the Ukrainian IDPs, the data collected in 2016 ranged from 21% to 32% and 47% of reported PTSD prevalence.^12, 19, 26^ By comparison, the current research found that 39% of IDPs obtained high levels of PTSD symptoms, whereas 55% met DSM-V criteria for PTSD diagnosis, that indicates an increase in prevalence. In 2016, anxiety was prevalent among 17% of IDPs,^12^ whereas, in our research, it increased to 49%. Indeed, a lack of available mental health care for Ukrainian IDPs has existed for several years; e.g., about 74% of IDPs who required mental health care in 2019 did not receive it.^27^

Although by April 2022, IDPs and refugees in Ukraine showed no significant difference in PTSD prevalence (about 31%),^2^ in our study, the refugees appeared to be the most vulnerable group by all measured variables in November 2022-January 2023. A recent paper reported anxiety symptoms among 51%, distress among 23%, and severe distress among 41% of adult refugees from Ukraine in Germany in May-August 2022.^10^ Current research shows similar anxiety levels: 52% of moderate and severe anxiety prevalence. However, we found 65% of moderate and 25% of severe stress as well as 47% and 63% of PTSD prevalence and severity. This finding might indicate a decrease in stress severity with a simultaneous extreme increase in PTSD development among Ukrainian refugees (compared to April 2022).^2^

In summary, all three groups evaluated require appropriate solutions from mental health care services in Ukraine and abroad, as well as further monitoring. At the time of the recent Russian invasion, the capacities of the Ukrainian mental health system insufficiently covered the needs of populations affected by the Crimea annexation and eight years of occupation in eastern Ukraine.^28^ The war escalation required urgent solutions to deal with previous and current gaps between the mental health issues of Ukrainians and relevant professional services.^29, 30^ With the support of international institutions, that is in high demand during the war,^25^ governmental and non-governmental institutions launched their projects and tools to provide Ukrainians with mental health services within the country and abroad.^31, 32^

Maintaining efforts for the sustainable development of the Ukrainian mental health service system and involving the best evidence-based practices to reduce trauma, mitigate stress and prevent further war-related consequences are needed.^31^

## LIMITATIONS

This study has several limitations related to the sample specifics and the research design. As for sample specifics, its gender distribution differs from other studies devoted to a similar purpose. For instance, recent papers on the mental health of Ukrainians in wartime reported the participation of 60%,^4^ 51%,^2^ 82% and 83% of women.^9, 20^ This difference we associate with the time of data collection and the sample specifics. Among the people located within Ukraine (NDPs and IDPs), the gender distribution of our respondents was 83-87%, close to the data collected by Chudzicka-Czupała and colleagues and Veldbrekht and Tavrovetska.^9, 20^ This might reflect the limited availability of men in wartime by November 2022 caused by imposition of martial law and general mobilization into the army. Furthermore, among surveyed refugees, females were 95% of cases, which we associate with banning men from leaving the country except for some categories of citizens. However, gender inequality might influence the study results, considering the recent finding that Ukrainian females in 2022 tended to have significantly higher levels of stress and anxiety.^9^ Second, although the sample size and specifics were calculated and composed under the purpose of a nationwide survey, we failed in providing equal distribution of respondent residences throughout all 24 oblasts of Ukraine and Crimea. Nevertheless, the sample size was large, and respondents represent all regions, i.e., western, eastern, central, northern, and eastern parts of Ukraine.

## CONCLUSIONS

War exposure and witnessing, along with the necessity to leave the region where they lived after 9-12 months of the 2022 Russian invasion, resulted in increased prevalence of stress, anxiety, and PTSD among Ukrainian adults. Refugees appeared to be the most vulnerable group, and IDPs felt much worse than NDPs. Representatives of all studied groups require further monitoring and adequate evidence-based interventions in mental health care aimed at reaching Ukrainians within the country and abroad.

## CONTRIBUTORS

OL contributed to the conceptualization of the study, the methodology, investigation, and acquired funding. OL, SB and VB curated the data. SB and VB did the formal analysis and validation of data. SB and OS contributed to data visualization OL, MV, SB and KB prepared the original draft. All authors contributed to reviewing and editing. OL supervised the study and had full access to raw data in the study. All authors have read and agreed to the published version of the manuscript.

## Data Availability

All data produced in the present work are contained in the manuscript

## ACKNOWLEDGEMENTS

This work was partially supported by grant from the Ministry of Education and Science of Ukraine (No 0123U101790). We also thank the many people who helped with data collection.

## COMPETING INTEREST STATEMENT

The authors have no competing interest to declare.

## Notes

### Competing Interest Statement

The authors have declared no competing interest.

### Author Declarations

Institutional Ethics Committee of Danylo Halytskyi National Medical University (protocol No 9, 22.11.2021)

